# Analysis of a Large Patient-Level Dataset to Predict Outcome of Treatment for Drug-Resistant Tuberculosis

**DOI:** 10.1101/2022.09.14.22279738

**Authors:** Qinlu Wang, Jingwen Gu, Andrei Gabrielian, Gabriel Rosenfeld, Mariam Quiñones, Darrell E. Hurt, Alex Rosenthal

## Abstract

**BACKGROUND:** Drug-resistant (DR) tuberculosis treatment is challenging and frequently leads to poor outcomes. An international collaboration, the National Institute of Allergy and Infectious Diseases (NIAID) TB Portals develops, maintains, and supports a multi-national database of tuberculosis cases, with an emphasis on drug-resistant tuberculosis. Patient records include clinical, radiological, genomic, and socioeconomic features. Establishing factors associated with unsuccessful treatment may help optimize treatment for the most challenging infections.

**METHODS:** Association analysis and machine learning algorithms were applied to identify important factors associated with treatment outcome and predict the outcome for three patient cohorts, selected by drug resistance level representing 1575 patients in total. The predicted probabilities of poor treatment outcome from models were calibrated as a risk score ranging from 0 to 100 corresponding to confidence level of the model for treatment outcome.

**RESULTS:** The features most associated with treatment success in all cohorts were body mass index (BMI), onset age, employment, education, smear-negative microscopy, and percent of abnormal volume in X-ray images, confirming previously reported findings, and identifying novel factors such as pathogen genomic markers.

**CONCLUSIONS:** The identified features might help in establishing high-risk patients at the time of admission for tuberculosis treatment. This study integrates clinical, radiological, and pathogen genomics into a patient risk model, a way of determining risk through the application of machine learning on real-world data.

## BACKGROUND

Tuberculosis (TB) is a communicable disease that is amongst the top 10 causes of death and the leading cause from a single infectious agent (1). With the Covid-19 pandemic’s ongoing global impact on healthcare systems, TB diagnostic and clinical management may experience challenges (2). The World Health Organization (WHO) released results of modeling work that if global TB case detection decreases by 25% over a period of 3 months (compared to level of detection pre-pandemic), an additional 190,000 TB deaths might occur (3). Thus, it is essential to identify the most vulnerable or hard-to-treat TB patients at the time of diagnosis. Identifying the most at-risk groups within the population may allow for better prophylactic measures and monitoring in advance, to prevent outbreaks of this highly contagious disease. While many tuberculosis databases or registries contain critical clinical information such as drug resistance status, time of diagnosis, sputum sampling, etc., it is rarer to find the integration of drug resistance with other important factors such as pathogen genomic status and social determinants of health.

Drug-resistant TB continues to be especially challenging to treat and control. Multidrug resistant TB (MDR-TB) is TB that is resistant to both rifampicin and isoniazid, two of the most widely used anti-TB drugs. Extensively drug-resistant TB (XDR-TB) is defined as MDR-TB plus resistance to at least one of the fluoroquinolones and one of the injectable agents used in MDR-TB treatment regimens (1). The latest global data show a treatment success rate of 85% for drug-susceptible TB, 56% for MDR-TB and 39% for extensively drug-resistant TB [1]. Given the lower success rate of treatment in drug resistance TB, the dynamics and nature of the disease show differences when comparing to drug sensitive TB and it is necessary to identify what clinical, genomic, or social determinants of health risk factors might be common or distinct between these disease types to highlight patient risk.

Prior studies have shown the importance of analyzing and interpreting the connections between clinical and social determinants of health. For example, clinical factors like culture-positivity at two months of treatment, history of treatment with second-line drugs were identified as risk factors of poor drug resistant tuberculosis treatment outcome in Eastern Europe and Central Asia (4). In the same study, the socio-economic factor of homelessness was also identified as correlating with poor outcome. Another study demonstrated a similar result where a combination of clinical factors such as MDR-TB patients with HIV, high smear grade, or a history of previous MDR-TB treatment were identified together with a socio-economic factor, malnutrition, as risk factors of poor treatment outcome (5). Another identified social determinants of health such as employment and education status along with clinical factors such as drug resistance status statistically associated and predictive of treatment failure (6). By analyzing a much larger dataset of clinical, radiological, and genomic features in a unique multi-national cohort of drug sensitive and drug resistant cases, our study extends earlier works that have investigated potential combinations of clinical and socio-economic factors impacting treatment outcome.

The TB Portals (https://tbportals.niaid.nih.gov/) is the largest, open-access, patient-centric database connecting clinical, socio-economic, pathogen genomics and patient radiological data from 16 countries (6-8). As part of the National Institute of Allergy and Infectious Diseases (NIAID) strategic plan for TB research and National Institute of Health (NIH) strategic plan for data science(9, 10), the resource is focused on how to translate real-world data collected from TB cases, primarily drug resistant, into actionable information for public health following the FAIR data principles. As part of this effort, the program considers researchers and public health experts with various backgrounds, providing ways to visualize, interact, and analyze the data. A request for data can be made by an investigator using the web-accessible data use agreement and application process, https://tbportals.niaid.nih.gov/download-data. The data can be accessed via API or directly downloaded to facilitate visualization, analysis, modeling, and other data science approaches. For users who prefer point and click interaction and analysis within a website, an ecosystem of tools appropriate for each type of data have been developed (8, 11, 12). The data and underlying suite of tools provides an unprecedented opportunity for public health researchers looking to understand the real-world impact of TB. For example, the program and its collaborators have demonstrated the applicability of TB Portals collection of data, publishing research on epidemiological aspects of tuberculosis, the evolutionary processes related to drug resistance, the factors involved with tuberculosis relapse versus reinfection, detailed analysis of various strains in sputum versus surgical samples of tuberculosis, analysis of radiological data to discover distinguishing features of drug resistant tuberculosis (7, 13, 14).

Here, we have analyzed real-world patients’ data (i.e. not intended as a representative epidemiological survey) as a retrospective case-control study using TB Portals data. Machine learning techniques were applied on 1575 TB cases with complete set of multi-domain data to predict the treatment outcome (either “cured” or “died”). Models were developed to each subgroup by drug resistance level in order to derive a case-level risk score to facilitate understanding of the relative contribution of clinical or social determinants of health factors towards the success of treatment. The insights provided by these models might assist in identifying the important risk factors that could inform public health policies and programs as part of a holistic, data-informed, and evidence-based approach. Moreover, this study is intended to demonstrate the potential of TB Portals for real-world studies and some of the important considerations of public health researchers interested in leveraging the resource.

## METHODS

### Data acquisition and initial processing

This analysis uses publicly shared, deidentified data from The Tuberculosis Data Exploration Portal (TB DEPOT) as of Mar 2021, which can be obtained by anyone who signs and agrees to a <u>data use agreement</u> (DUA). Only cases resulting in death (died, negative outcome) or recovery (cured, positive outcome) at the end of the treatment with available BMI and age measures were included. There were 1575 subjects with features of interest, including 299 drug-sensitive (DS) TB patients, 883 MDR-TB patients, and 393 XDR-TB patients. The clinically reported type of resistance was used throughout the study. These patients came from multiple countries spanning the consortium (Table 1 and Table S1). Categorical feature levels with limited samples were combined: for gender, female and other were combined as non-male; for education, basic school and no education were combined as basic school or lower, College (bachelor) and Higher (university) were combined as college or higher; for employment, Homemaker and Self-employed were combined as Homemaker or self-employed. Missing categories (“Not reported”, “NA”, blank) were combined into a single missing category for any categorical data. A case was considered as resistant if the pathogen showed resistance in any of First-line Drug Line Probe Assay, Second-line Drug Line Probe Assay, Solid medium Lowenstein, BACTEC MGIT 960, or GeneXpert MTB/RIF (Xpert) tests. Certain variables from lung images are subdivided by sextant of the lung corresponding to upper, middle, lower sextant of right or left lung. A case might have multiple lung sextants involving a feature from an available image or multiple images per case. In the following analysis, the levels are combined into “Upper sextant - Yes” or “No”, “Middle or Lower sextant - Yes” or “No” to indicate the combination of features from available image data.

**Table 1.**
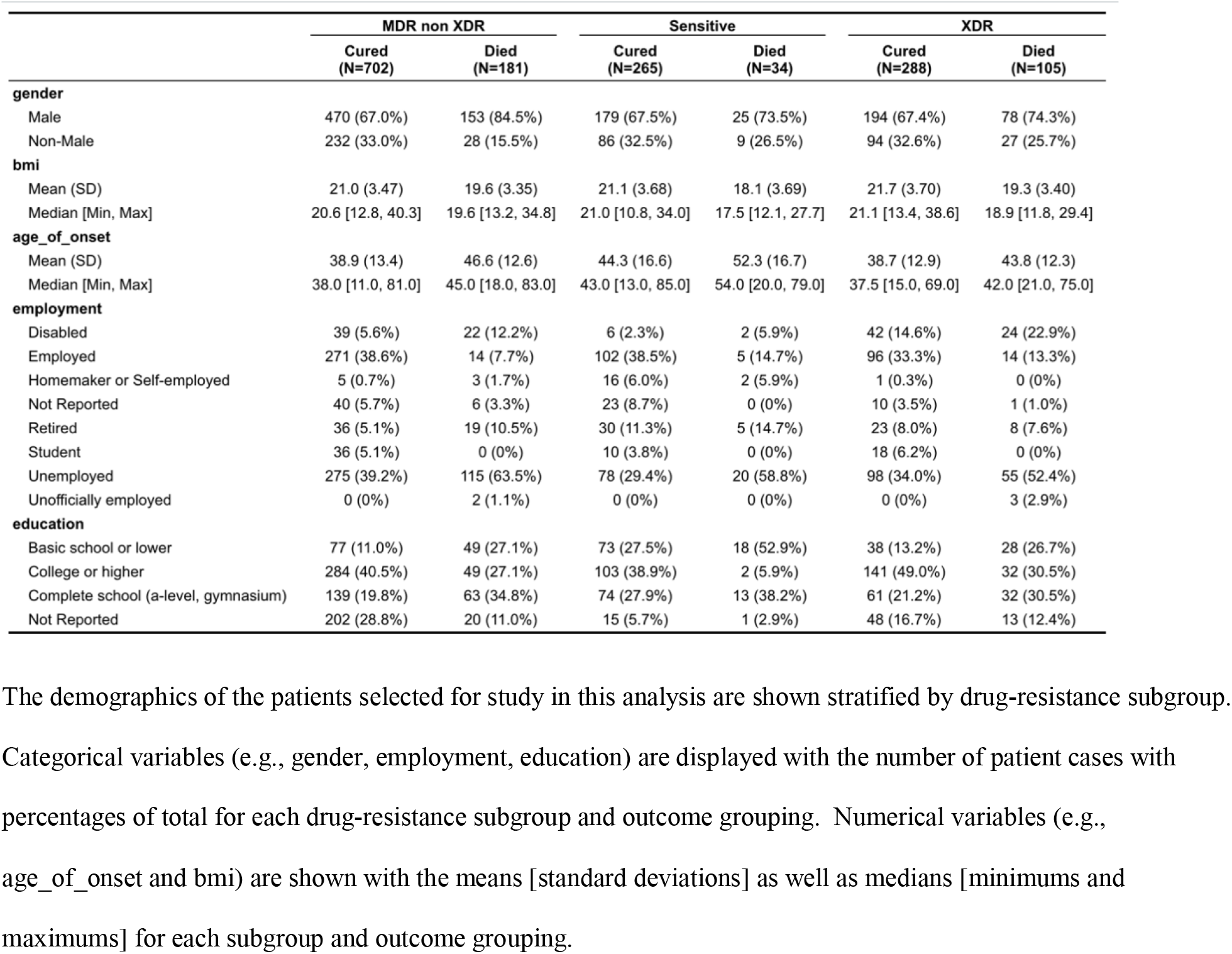
Demographics of Different Drug-Sensitive and Drug-Resistant Subgroups

|  | MDR non XDR |  | Sensitive |  | XDR |  |
| --- | --- | --- | --- | --- | --- | --- |
|  | Cured (N=702) | Died (N=181) | Cured (N=265) | Died (N=34) | Cured (N=288) | Died (N=105) |
| **gender** |  |  |  |  |  |  |
| Male | 470 (67.0%) | 153 (84.5%) | 179 (67.5%) | 25 (73.5%) | 194 (67.4%) | 78 (74.3%) |
| Non-Male | 232 (33.0%) | 28 (15.5%) | 86 (32.5%) | 9 (26.5%) | 94 (32.6%) | 27 (25.7%) |
| **bmi** |  |  |  |  |  |  |
| Mean (SD) | 21.0 (3.47) | 19.6 (3.35) | 21.1 (3.68) | 18.1 (3.69) | 21.7 (3.70) | 19.3 (3.40) |
| Median [Min, Max] | 20.6 [12.8, 40.3] | 19.6 [13.2, 34.8] | 21.0 [10.8, 34.0] | 17.5 [12.1, 27.7] | 21.1 [13.4, 38.6] | 18.9 [11.8, 29.4] |
| **age_of_onset** |  |  |  |  |  |  |
| Mean (SD) | 38.9 (13.4) | 46.6 (12.6) | 44.3 (16.6) | 52.3 (16.7) | 38.7 (12.9) | 43.8 (12.3) |
| Median [Min, Max] | 38.0 [11.0, 81.0] | 45.0 [18.0, 83.0] | 43.0 [13.0, 85.0] | 54.0 [20.0, 79.0] | 37.5 [15.0, 69.0] | 42.0 [21.0, 75.0] |
| **employment** |  |  |  |  |  |  |
| Disabled | 39 (5.6%) | 22 (12.2%) | 6 (2.3%) | 2 (5.9%) | 42 (14.6%) | 24 (22.9%) |
| Employed | 271 (38.6%) | 14 (7.7%) | 102 (38.5%) | 5 (14.7%) | 96 (33.3%) | 14 (13.3%) |
| Homemaker or Self-employed | 5 (0.7%) | 3 (1.7%) | 16 (6.0%) | 2 (5.9%) | 1 (0.3%) | 0 (0%) |
| Not Reported | 40 (5.7%) | 6 (3.3%) | 23 (8.7%) | 0 (0%) | 10 (3.5%) | 1 (1.0%) |
| Retired | 36 (5.1%) | 19 (10.5%) | 30 (11.3%) | 5 (14.7%) | 23 (8.0%) | 8 (7.6%) |
| Student | 36 (5.1%) | 0 (0%) | 10 (3.8%) | 0 (0%) | 18 (6.2%) | 0 (0%) |
| Unemployed | 275 (39.2%) | 115 (63.5%) | 78 (29.4%) | 20 (58.8%) | 98 (34.0%) | 55 (52.4%) |
| Unofficially employed | 0 (0%) | 2 (1.1%) | 0 (0%) | 0 (0%) | 0 (0%) | 3 (2.9%) |
| **education** |  |  |  |  |  |  |
| Basic school or lower | 77 (11.0%) | 49 (27.1%) | 73 (27.5%) | 18 (52.9%) | 38 (13.2%) | 28 (26.7%) |
| College or higher | 284 (40.5%) | 49 (27.1%) | 103 (38.9%) | 2 (5.9%) | 141 (49.0%) | 32 (30.5%) |
| Complete school (a-level, gymnasium) | 139 (19.8%) | 63 (34.8%) | 74 (27.9%) | 13 (38.2%) | 61 (21.2%) | 32 (30.5%) |
| Not Reported | 202 (28.8%) | 20 (11.0%) | 15 (5.7%) | 1 (2.9%) | 48 (16.7%) | 13 (12.4%) |

The demographics of the patients selected for study in this analysis are shown stratified by drug-resistance subgroup. Categorical variables (e.g., gender, employment, education) are displayed with the number of patient cases with percentages of total for each drug-resistance subgroup and outcome grouping. Numerical variables (e.g., age\_of\_onset and bmi) are shown with the means [standard deviations] as well as medians [minimums and maximums] for each subgroup and outcome grouping.

### Association Analysis

Measure of effect size in ANOVA (analysis of variance) quantifies degree of association between an effect and a continuous dependent variable. Eta squared (*η*^2^) was used to measure size of observed effect with age and BMI. Cohen defined small, medium, and large effects as Eta squared values of 0.01, 0.06, 0.14 respectively (15). Variables with statistically significant difference in the mean of age or BMI by drug-sensitive and drug-resistant subgroups were reported.

Uncertainty coefficient (UC) was applied to measure the association between pairs of variables (e.g., gender and other non-continuous variables). UC represents a percent reduction in error when predicting dependent variable from independent variable. When UC is 0, the independent variable lacks information to predict the dependent variable (16). The association of variables that were statistically significant with outcome by Fisher exact test were reported. The UC association analysis excluded missing values.

### Prediction of Treatment Outcome

Categorical covariates with more than 2 levels were encoded into binary variables. Features were standardized by min-max normalization. Features with highest UC with treatment outcome were selected for each drug-sensitive and drug-resistant TB subgroup respectively. Seventy percent of data were selected as a training dataset and the remainder were used as an independent testing dataset. Machine learning algorithms, Logistic Regression, Random Forest, Support Vector Machine, and XGBoost, were trained on the training set to predict treatment outcome. Repeated grid search 3-fold cross validation was used for parameter tuning. Features with more than 75% missing data or less than 1% variation were removed. Synthetic Minority Oversampling Technique (SMOTE) was used to oversample the minority class, of “died”. Area Under the Receiver Operating Characteristics Curve (AUROC) and Area Under the Precision and Recall Curve (PRAUC) were used to evaluate model performance on the testing dataset. Feature importance was generated for the model with highest AUROC and PRAUC. Model coefficients and Shapely Additive Explanations (ShAP) indicate feature importance for Logistic Regression and tree-based models respectively.

### Risk Score

The predicted probabilities of poor treatment outcome from models for each drug-sensitive or drug-resistant TB patient subgroup were calibrated as a risk score ranging from 0 to 100 corresponding to confidence level of the model for treatment outcome. The risk scores of the training dataset were calculated by out-of-bag cross-validation while the risk scores of the testing dataset were calculated by the best model trained on the training dataset.

### Statistical Software

The data cleaning, inferential statistics, and association analysis were done using R statistical software version 4.0.2 (RStudio version 1.2.5033) (17) with packages fastDummies, dplyr, ggplot2, finalfit. The predictive modeling was done using Python 3 (18) with packages numpy, pandas, matplotlib, sklearn, random, and xgboost.

## RESULTS

### Demographics of identified cohort stratified by drug-resistance

Given our focus on predicting the outcome within the specific drug resistance subtypes and ensuring that the identified factors are generalizable, we first assessed the resistance status of the selected cohort of patients with regards to outcome. The treatment success rate for DS-TB cases was high (88.6%), while it was lower for MDR-TB treatment (79.5%), and even lower for XDR-TB treatment (73.3%) (Table S2). While the rates of treatment success outcome correlate with the ones from the WHO report in 2019 (1), the absolute rates are distinct because only the most definitive outcomes of died and cured patients were considered in this analysis while the WHO report considered died, cured, treatment failed, and lost-to-follow-up patients. Importantly, there was a significant difference of success rates among these drug-sensitive and drug-resistant subgroups (p < 0.0001, Chi-Square Test) supporting the strategy of stratifying by resistance subgroup in the machine learning analyses.

### Association Analysis

With the diverse demographic, clinical, radiological, microbiological, and genomic domains of data available in TB Portals, we studied the most significant inter-domain relationships using association analysis. The goal was to explore these inter-domain relationships prior to including these variables in the modeling. This approach attempts to identify any potential biases as well as highlight significant associations between types of data (e.g., pathogen genomics and imaging) taking advantage of the patient-centric, multi-domain nature of the TB portals resource. As these domains of data have not been combined previously, there was an opportunity for discovery as well as comparing known associations from prior studies.

Among the top variables associated with outcome in DS, MDR, and XDR TB were imaging related pathologies like nodule, cavity, and fibrosis as well as overall area of abnormality. Moreover, non-imaging features including social determinants of health like employment and education status are present across resistance types. The complete list of these factors is in Table 2. The heatmaps of the most important variables associated with treatment outcome in MDR-TB and XDR-TB groups are visualized

**Table 2.**
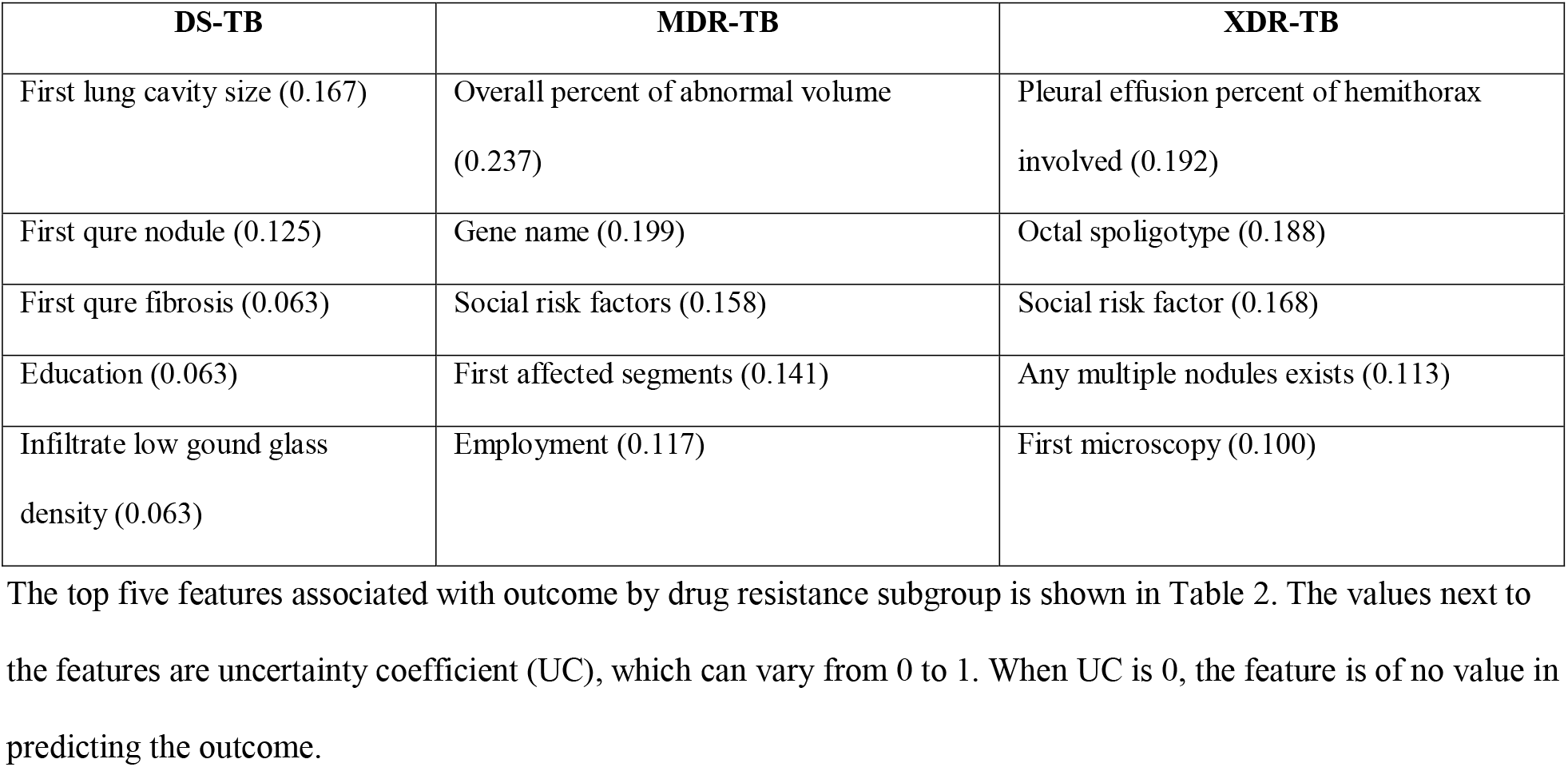
Top variables associated with Outcome by UC

(Figure 1). In the XDR-TB group, UC with outcome were between 0.090 and 0.192. The top associated feature *Pleural effusion percent of hemithorax* was in the same cluster with outcome. In MDR-TB group, UC with outcome were between 0.07 and 0.237. For full code and complete overview of the association analysis pipeline, please refer to our GitHub repository (https://github.com/niaid/tb-portals-association-and-prediction).

**Figure 1.**
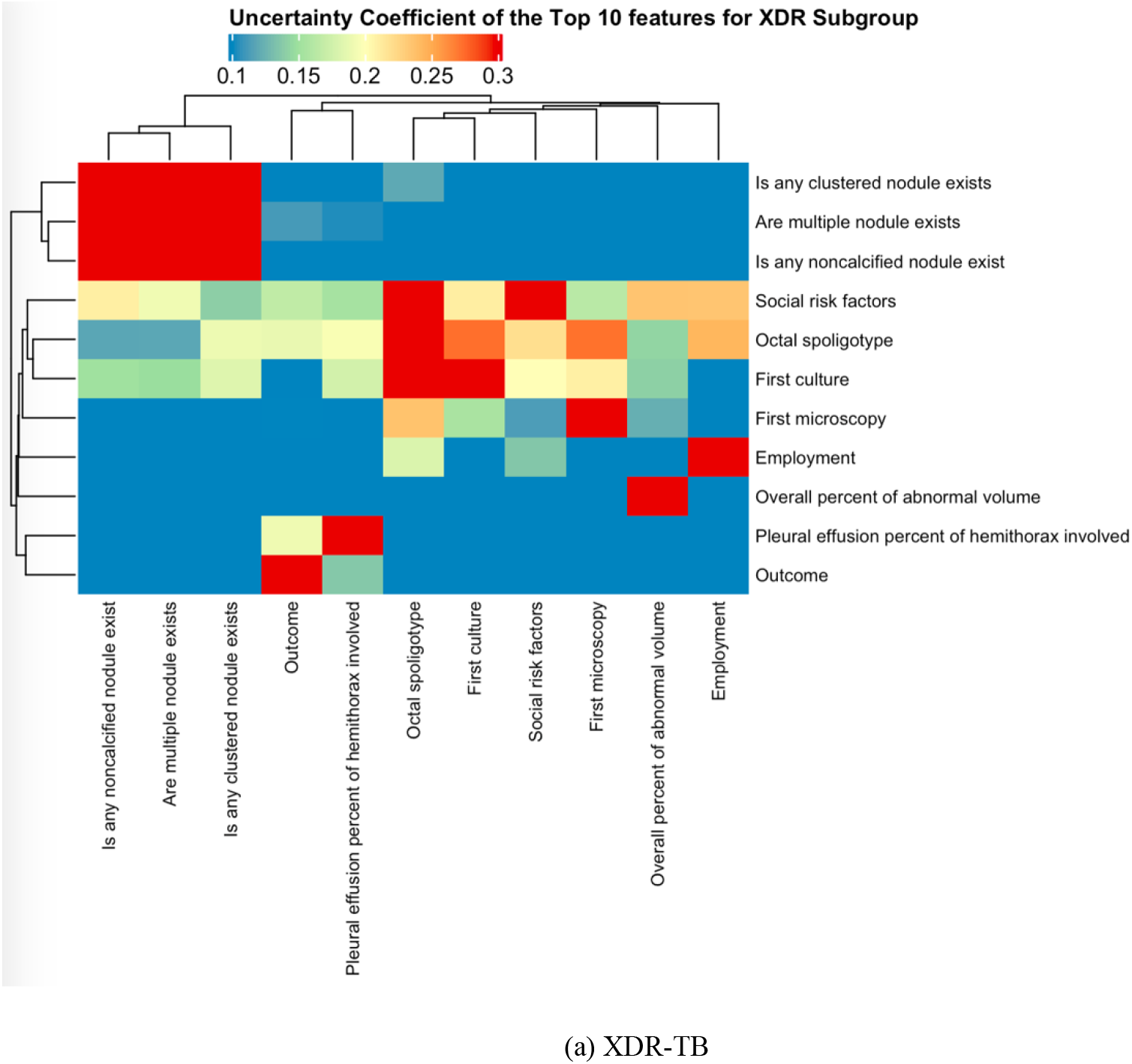
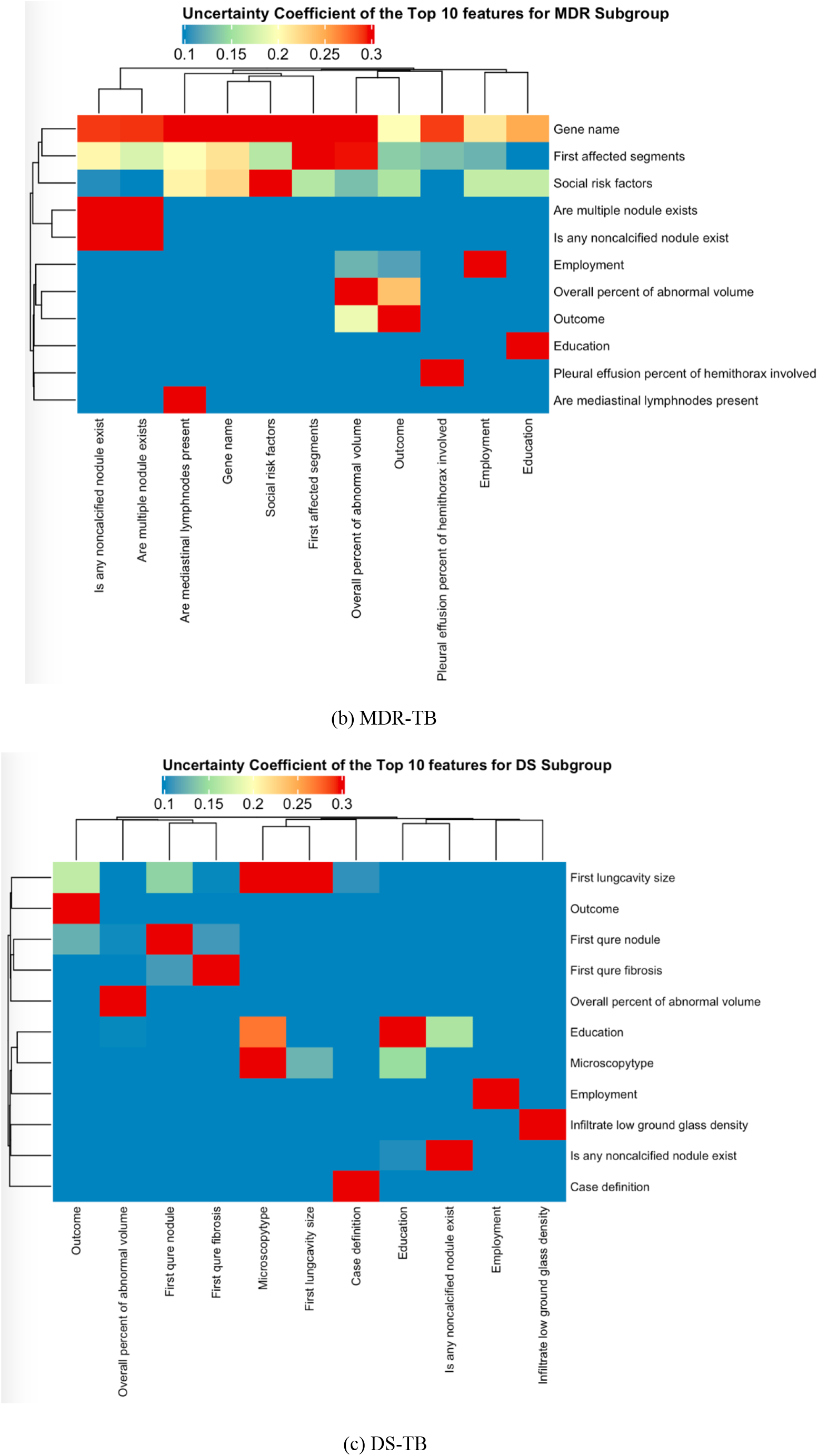
Heatmap of Top Ten Features associated with Treatment Outcome. The top ten features associated with Treatment Outcome are shown in relation to each other with a higher uncertainty coefficient (UC) indicating a stronger association between sets of features. In A), the relationship by UC for the top 10 features for XDR-TB (N = 430) are shown while in B), the relationship is shown for MDR-TB (N = 1019) and C), the relationship for DS-TB (N = 811). In both, the sets of features are clustered by hierarchical clustering to show clusters of features showing associations.

### Modeling and Prediction

We were able to obtain reasonable predictive capacity across a variety of algorithms; however, we did observe certain models performing better depending upon the specific resistance subgroup. The performance of different predictive models was compared (Table 3 and Figure 2). XGBoost outperformed other models in the DS-TB patient subgroup (AUROC = 0.8188, PRAUC = 0.5836); Logistic Regression with Lasso regularization outperformed other models in the MDR-TB patient subgroup (AUROC = 0.8353, PRAUC = 0.6037); in XDR-TB patient subgroup, Random Forest outperformed other models (AUROC = 0.8665, PRAUC = 0.7260). The top features associated with treatment outcome with the highest feature importance were selected in DS-TB, MDR-TB, XDR-TB patient subgroups (Figure 3).

**Table 3.** Comparison of model performance

|  | **DS-TB (N=299)** |  | **MDR-TB (N=883)** |  | **XDR-TB (N=393)** |  |
| --- | --- | --- | --- | --- | --- | --- |
| **Models** | **AUROC** | **PRAUC** | **AUROC** | **PRAUC** | **AUROC** | **PRAUC** |
| Logistic Regression | 0.8688 | 0.3758 | 0.8353 | 0.6037 | 0.8438 | 0.7234 |
| Random Forest | 0.8050 | 0.4144 | 0.8138 | 0.5562 | 0.8665 | 0.7260 |
| Support Vector Machine | 0.8288 | 0.3258 | 0.8180 | 0.5572 | 0.8020 | 0.6523 |
| XGBoost | 0.8188 | 0.5836 | 0.8198 | 0.5382 | 0.8230 | 0.6846 |

The performance of different predictive models in Area Under the Receiver Operating Characteristics Curve (AUROC) and Area Under the Precision and Recall Curve (PRAUC) are shown stratified by drug-resistance subgroup. The models with the highest PRAUC are highlighted.

**Figure 2.**
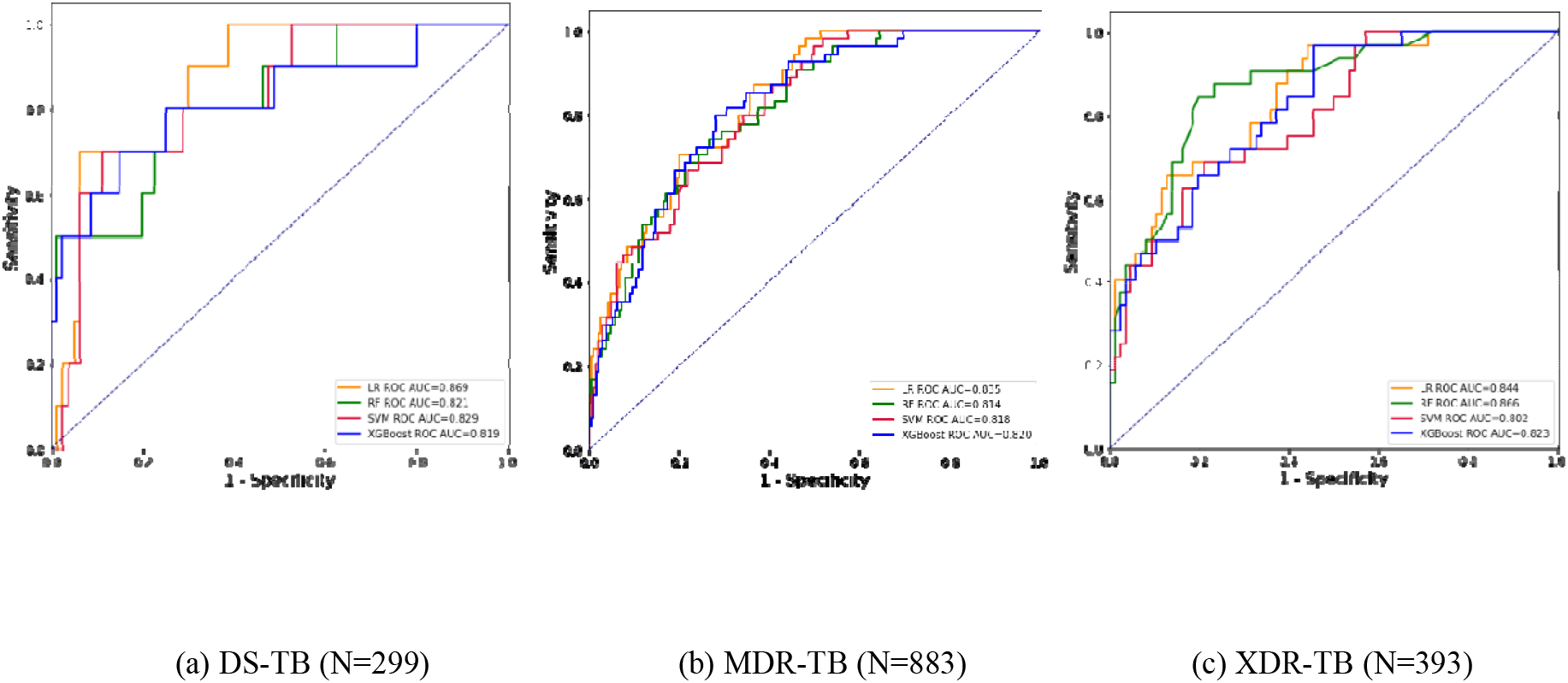
ROC Curves of Drug-Sensitive and Drug-Resistant Subgroups The receiver operating characteristic (ROC) Curves of different predictive models are shown stratified by drug-resistance subgroup. The Area Under the Receiver Operating Characteristics Curve (AUROC) for a given curve is the area beneath it. The AUROC values are listed in the legends.

**Figure 3.**
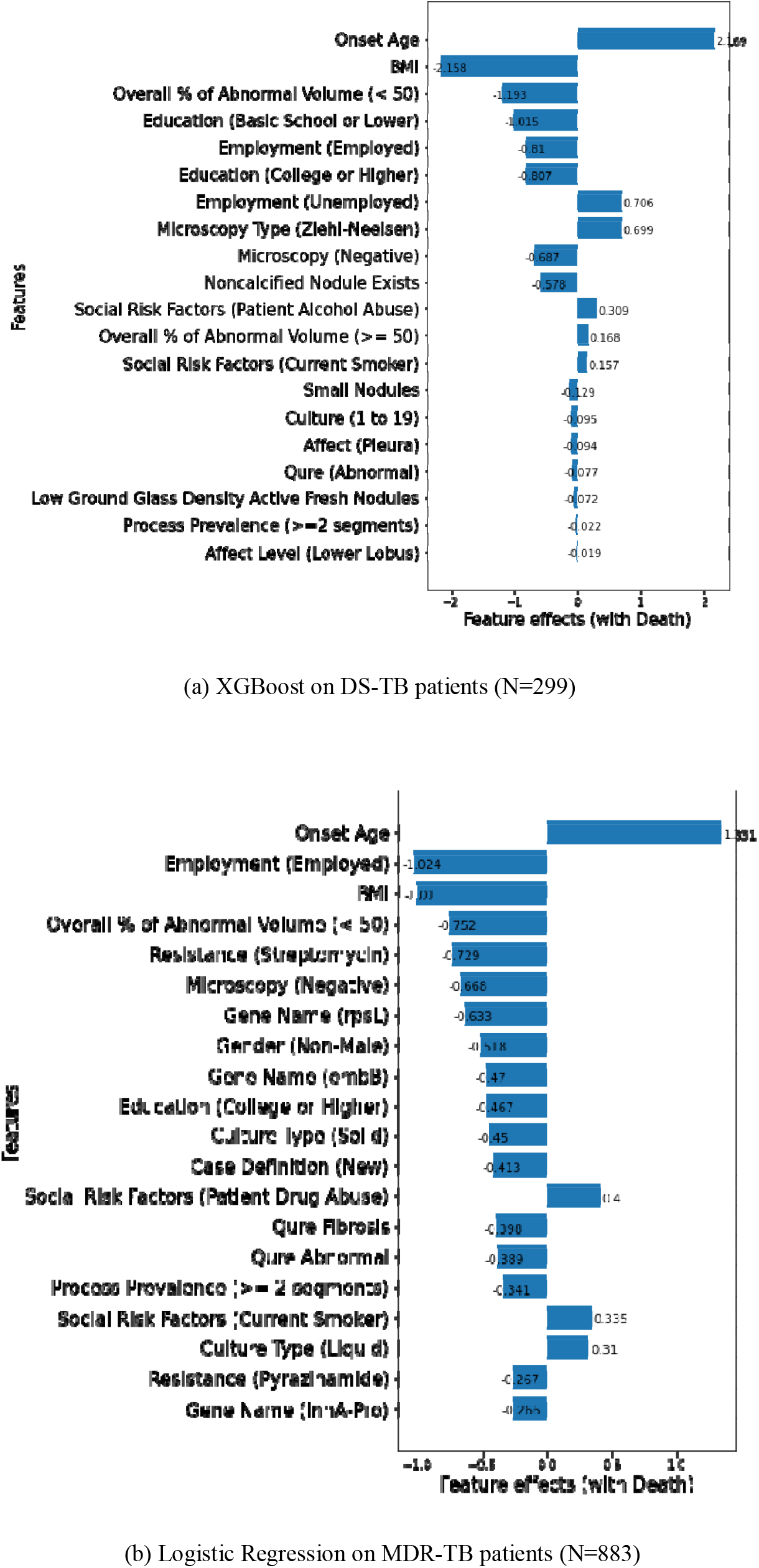
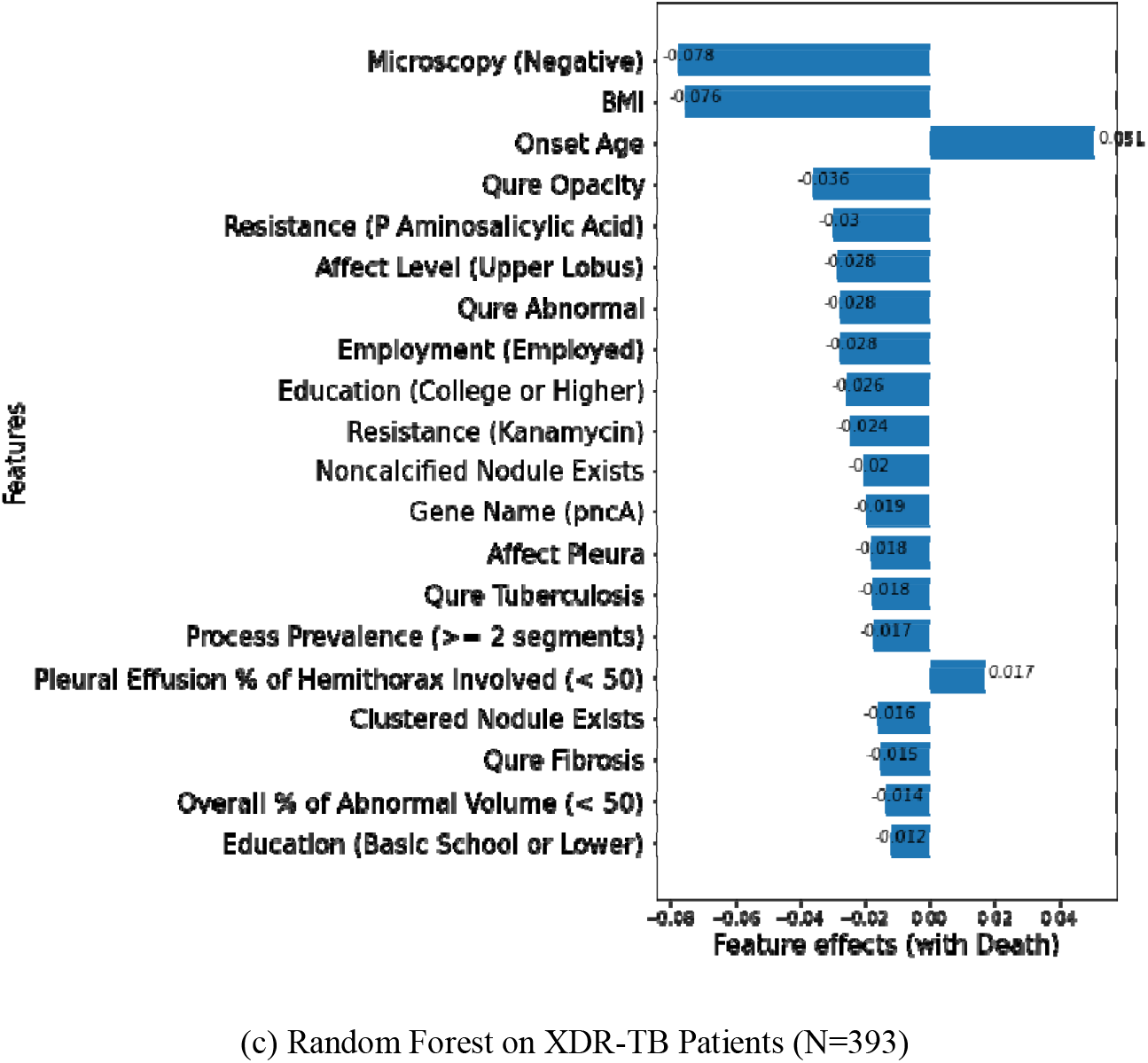
Feature Importance of top 20 features from the Best Models For Logistic Regression, the plot shows that normalized features with positive feature effects increase the odds of outcome of death compared to cured whereas normalized features with negative feature effects show the opposite. For tree-based models, the plot shows Shapley values and features with larger absolute Shapley values are more important contributors to the prediction.

This could explain why specific models performed better in particular resistance groups since the variables may show non-linear or linear dependencies with the outcome of interest. The boxplot of top features was generated for each drug-sensitive and drug-resistant subgroup (Figure S1). The frequency tables and univariate odds ratios between top features with treatment outcome were listed in Table S5.

### Risk Scores

The risk scores were derived from the predictive probabilities of all-cause mortality from the best predictive model for each subgroup respectively. There is a statistically significant difference in risk scores between cured and died outcome for TB patients by Two-sample t-test (Figure S2). For most cases, risks aligned with the expected outcome (Figure S3). In public health and clinical practice, it is important to identify and highlight situations where predicted risks do not align with expected outcome due to potential biases in data collection or other causes. We examined these on a case-by-case basis to account for potential impacts of social determinants of health or other factors on predicted outcome. Our risk analysis provides new understanding of the mechanisms of risk that can inform future clinical study and healthcare policy specific to important subtypes of TB (e.g., drug-sensitive versus drug-resistant).

## DISCUSSION

TB Portals presents the largest, publicly available real-world dataset of tuberculosis cases to understand the important clinical and socio-economic features of the case that predict treatment outcome. We analyzed the data from TB Portals and identified factors spanning the distinct domains of information including clinical and socio-economic features to discover associations and interactions between these domains that might impact the probability of a poor treatment outcome. We confirmed prior risk factors identified in earlier studies while extending the analysis to include stratification of drug resistance status, which allows for a more accurate understanding of the common or divergent factors across subgroups. Six features were associated with treatment outcome in all three subgroups: BMI, onset age, employment, education (college or higher), smear-negative microscopy, and overall percent of abnormal lung volume as determined by a radiologist in X-ray images (less than 50 percent). We confirmed in a large, stratified analysis several previously reported clinical as well as socio-economic factors like BMI, age, microscopy, and imaging as important towards the probability of successful treatment outcome.

BMI was found to be the most predictive feature inversely associated with all-cause mortality, consistent with population-based cohort studies by Hsien-Ho, et al. (19). Employment is usually associated with better healthcare outcomes as it is indicative of higher income, health insurance coverage, and socio-economic status. College or higher education as well as employment status is inversely associated with all-cause mortality. Onset age is a predictor of all-cause mortality consistent with an earlier study finding that older age associated with treatment failure (20). Smear microscopy is commonly used as a part of the primary diagnostic protocol and monitoring of treatment efficacy of TB in many low- or medium-income countries. In our analysis, we found that negative smear microscopy was associated with successful treatment, which is consistent with clinical guidelines indicating treatment efficacy. Aside from critical demographic and social determinants of health factors, our analysis identified several radiological (overall abnormal lung volume, presences of nodules, upper lobe involvement, etc.) as well as pathogen genomic features (octal spilogotype and genomic variants in select resistance genes). These distinct domains of data have not frequently been analyzed together; and we believe it is necessary to consider them together to understand the holistic nature of the disease and treatment.

There are some caveats that need to be carefully considered. The temporal dynamics in a TB case are complex and require an expanded dataset with information captured at distinct timepoints of the treatment, which is also available from TB Portals (https://analytic.tbportals.niaid.nih.gov/index.html). The aforementioned API as well as data sharing website describing the data model (https://datasharing.tbportals.niaid.nih.gov/#/about-the-data) present an opportunity to assess each case temporally through relational organization of the timing of important events (imaging, treatment, culture, and microscopy) in number of days from the earliest registration date in the record. The presented results in this study focus on the case level summary and we plan to expand these analyses in the future to account for temporal dynamics in the case. For example, we summarized information about radiologist reported lung pathology that is captured across one or more images and involving segments within the lung into categories (e.g., “Upper sextant - Yes” or “No”, “Middle or Lower sextant - Yes” or “No” to indicate the combination of features from available image data). We focused on understanding the factors associated with cured and died outcomes to examine the predictors of the most definitive outcomes; however, we plan to follow up with additional analyses that examine other outcomes since competing risks might overlap between end points such as all-cause-mortality or treatment failure.

TB portals is a real-world data resource focusing on the most challenging TB cases and the programmatic priorities of participating clinical centers and so it is enriched in highly-drug resistant cases as a natural history study. The result from any machine learning and association study should not be considered for making actionable clinical decisions until a clinical study or clinical trial demonstrates efficacy of an intervention. Understanding the importance of bias or areas of failure in model performance is also important for the application of machine learning in public health. We explored some of these dynamics using Shapely Additive Explanations (SHAP) force plots (Figure S3) in a case-by-case basis for outlier or unexpected predictions to highlight situations in which the predictions from our models conflict with expected observations. Given that these are real-world data, our findings need to be interpreted cautiously due to the potential of confounding from observed or unobserved variables; however, the analysis of cross-domain information can provide valuable insight for future translational medicine efforts and study. We plan to periodically update this analysis with new information and cases as data become available from the rapid growth of the TB Portals resource.

## Data Availability

The TB portals program necessitates all users of the data sign a DUA before access to the underlying, de-identified clinical data is provided and the data can be requested at the following URL (https://tbportals.niaid.nih.gov/download-data). Therefore, this study provides code used in the analysis without the underlying raw data (https://github.com/niaid/tb-portals-association-and-prediction) in compliance with the DUA.

## LIST OF ABBREVIATIONS

DR: Drug resistant
BMI: Body mass index
NIAID: National Institute of Allergy and Infectious Diseases
NIH: National Institute of Health
TB: Tuberculosis
MDR-TB: Multidrug resistant TB
XDR-TB: Extensively drug-resistant TB
WHO: World Health Organization
TB DEPOT: Tuberculosis Data Exploration Portal
DUA: Data usage agreement
DS: Drug-sensitive
ANOVA: Analysis of variance
UC: Uncertainty coefficient
SMOTE: Synthetic Minority Oversampling Technique
AUROC: Area Under the Receiver Operating Characteristics Curve
PRAUC: Area Under the Precision and Recall Curve
SHAP: Shapely Additive Explanations

## DECLARATIONS

### ETHICS APPROVAL AND CONSENT TO PARTICIPATE

The data is provided by the TB Portals program stripped of all identifiers as a de-identified dataset according to their data use agreement (https://tbportals.niaid.nih.gov/pdf/TB-Portals-Data-Use-Agreement.pdf). All methods were carried out in accordance with relevant guidelines and regulations.

### CONSENT FOR PUBLICATION

Not applicable

### AVAILABILITY OF DATA AND MATERIALS

The deidentified datasets are publicly available in the Tuberculosis Data Exploration Portal (TB DEPOT), https://depot.tbportals.niaid.nih.gov/#/home.

### AUTHORS’ CONTRIBUTIONS

QW applied machine learning algorithms to predict the outcome of tuberculosis patients and wrote the manuscript. JG conducted association analysis to identify important factors associated with treatment outcome. AG initialized the project, helped interpreting the results clinically and writing the conclusion. GR edited the manuscript and assisted with submission. MQ, DH, AR provided guidance on the project and revised the manuscript. All authors read and approved the final manuscript.

## ACKNOWLEDGEMENTS

We thank Kurt Wollenberg, PhD from NIAID for his suggestions and support on this project; Alina Grinev, MD, MSBM from NIAID for managing this project; Alyssa Long, BS from NIAID for accessing the TB Portals database.

