## Supplementary Materials for "Analysis of a Large Patient-Level Dataset to Predict Outcome of Treatment for Drug-Resistant Tuberculosis"

Table S1 - Demographics of Different Drug-Sensitive and Drug-Resistant Subgroups

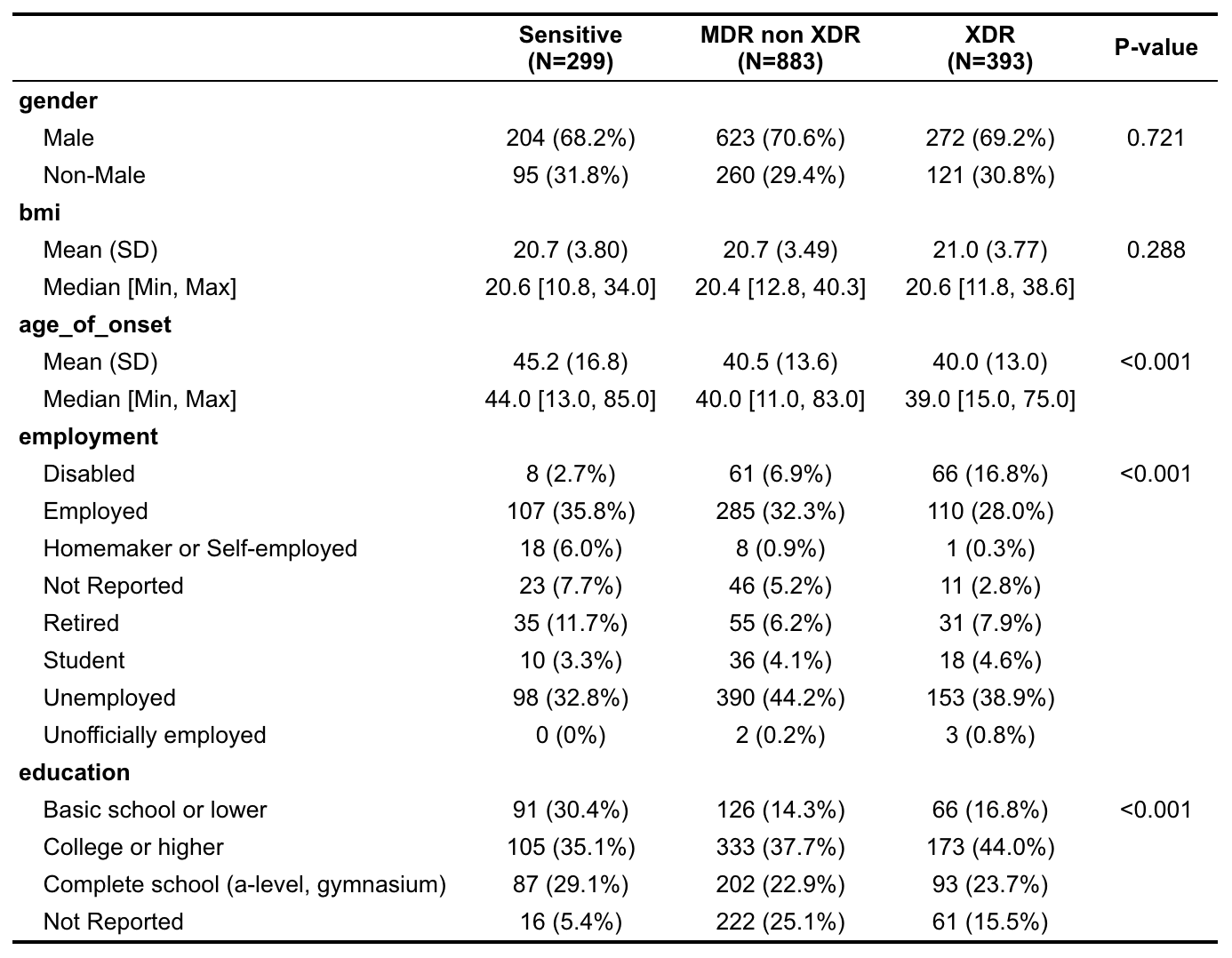

The demographics of the patients selected for study in this analysis are shown stratified by drug-resistance subgroup. Categorical variables (e.g. gender, employment, education) are displayed with the number of patient cases with percentages of total for each drug-resistance subgroup and outcome grouping. Numerical variables (e.g., age_of_onset and bmi) are shown with the means [standard deviations] as well as medians [minimums and maximums] for each subgroup and outcome grouping. Chi-square test evaluate the relationship between demographic factors with drug-resistance

Table S2 – Treatment Success Rates of Different Drug-Sensitive and Drug-Resistant Subgroups

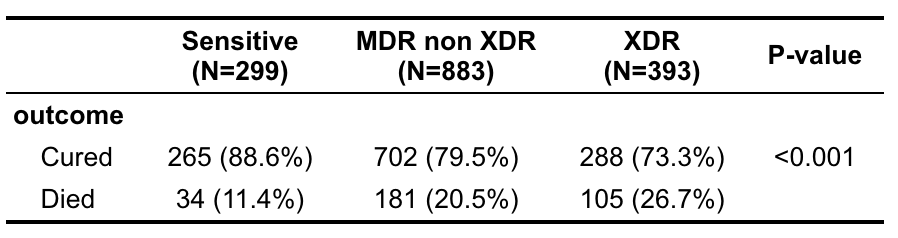

The counts of cured cases and died cases with the percentages of total for each drug-resistance subgroup and outcome grouping. There was a significant difference of success rates among these drug-sensitive and drug-resistant subgroups (p < 0.0001, Chi-Square Test).

Table S3 – Top variables associated with Age by effect size in ANOVA

| **Sensitive TB** | **MDR TB** | **XDR TB** |
| --- | --- | --- |
| Octal spoligotype (0.313) | Regimen drug (0.466) | Regimen drug (0.683) |
| Employment (0.309) | First regimen (0.351) | First Regimen (0.549) |
| First nodicalcinatum (0.272) | Employment (0.294) | Employment (0.353) |
| First affected segments (0.151) | Octal spoligotype (0.129) | Gene name (0.173) |
| First affected level (0.132) | Gene name (0.124) | First affected segments (0.164) |

Table S4 – Top variables associated with BMI by Effect size in ANOVA

| **Sensitive TB** | **MDR TB** | **XDR TB** |
| --- | --- | --- |
| Octal spoligotype (0.510) | Regimen drug (0.453) | Regimen drug (0.786) |
| lineage (0.249) | First regimen (0.346) | First regimen (0.638) |
| Education (0.142) | Gene name (0.192) | Gene name (0.301) |
| Overall percent of abnormal volume (0.140) | Octal spoligotype (0.172) | Social risk factor (0.154) |
| First affected segments (0.113) | Social risk factors (0.091) | Octal spoligotype (0.147) |

The top associated variables with age and BMI by resistance subgroup is shown in Table S3 and S4. The strength of the observed effect is measured by Eta-square, which ranges between 0 and 1. The standard for small, medium, large effect are 0.01, 0.06, 0.14 respectively by Cohen [10].

Table S5 - Frequency Table and Univariate Odds Ratio for Top 10 Selected Features

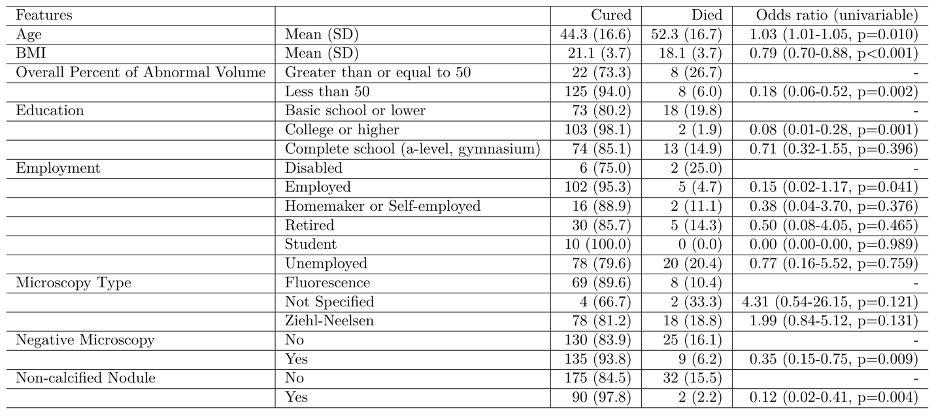

(a) DS-TB (N=299)

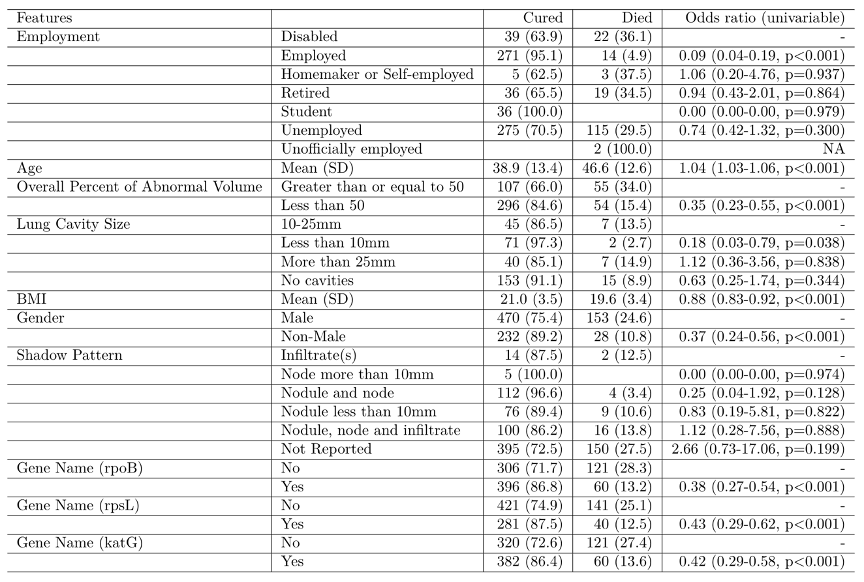

(b) MDR-TB (N=883)

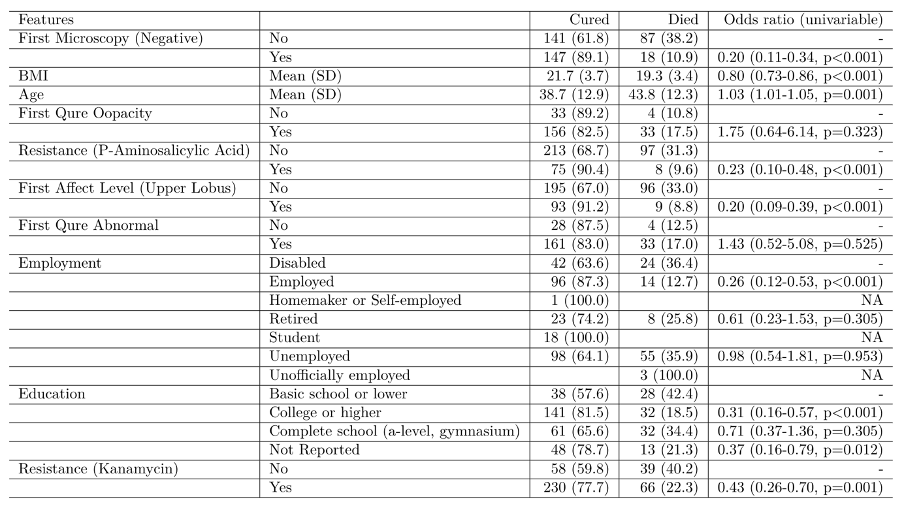

(c) XDR-TB (N=393)

Frequency Table and Univariate Odds Ratio for Top 10 Selected Features are shown stratified by drug-resistance subgroup. Chi-squared tests with a continuity correction for categorical variables evaluates the independence between features and treatment outcome. NA means limited sample size with no variation.

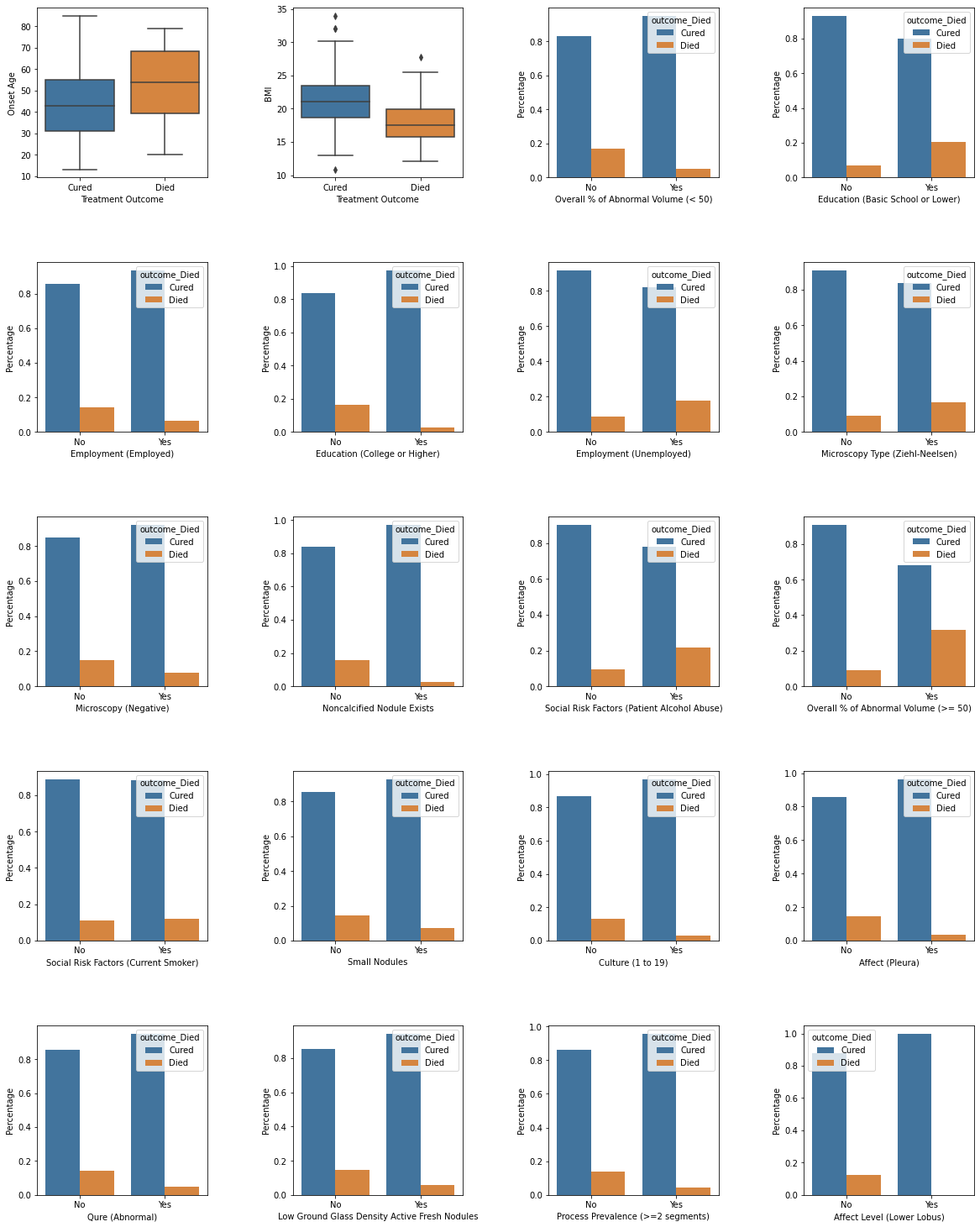

Figure S1 (a) - Top 20 Features Associated with Treatment Outcome – DS-TB

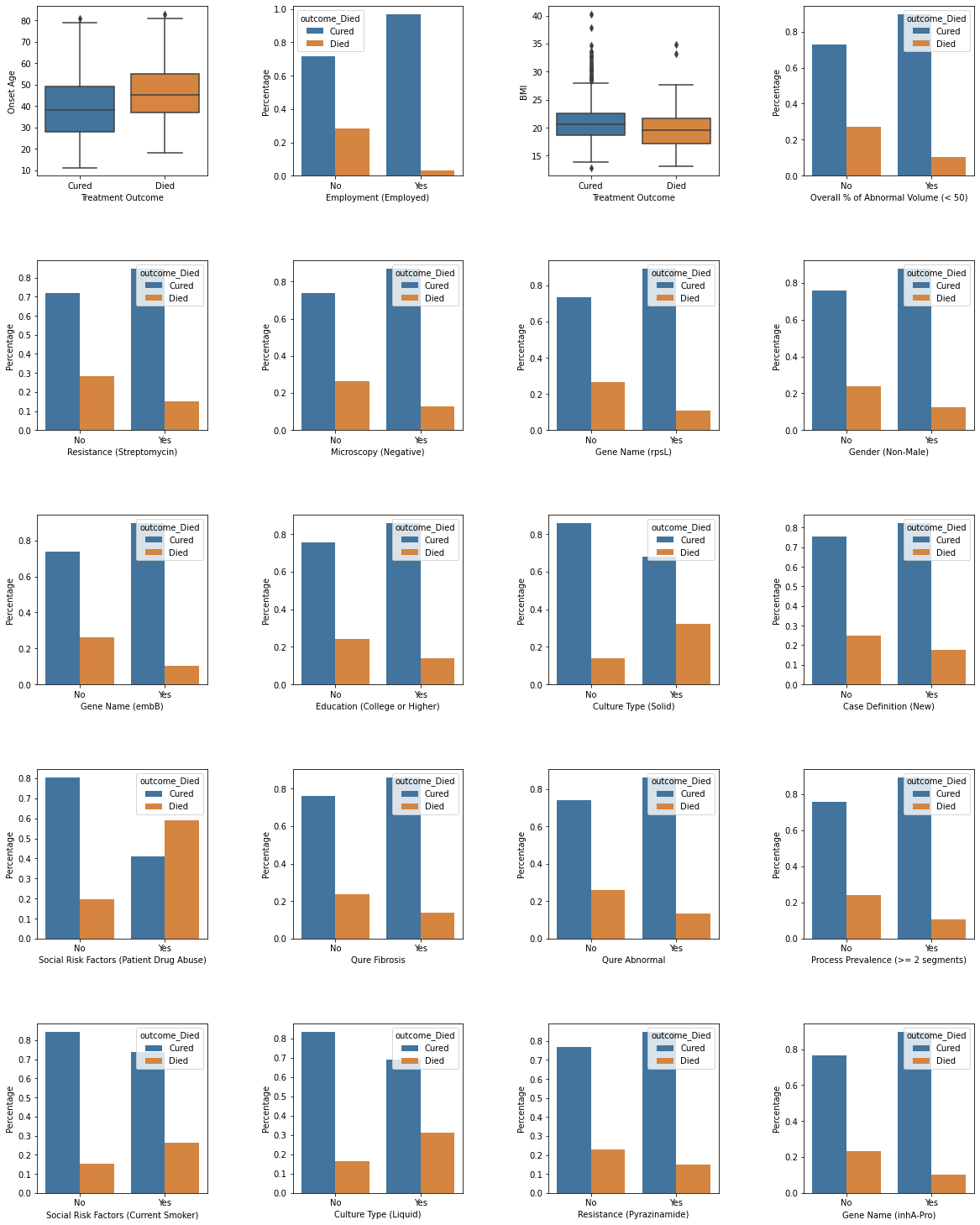

Figure S1 (b) - Top 20 Features Associated with Treatment Outcome – MDR TB

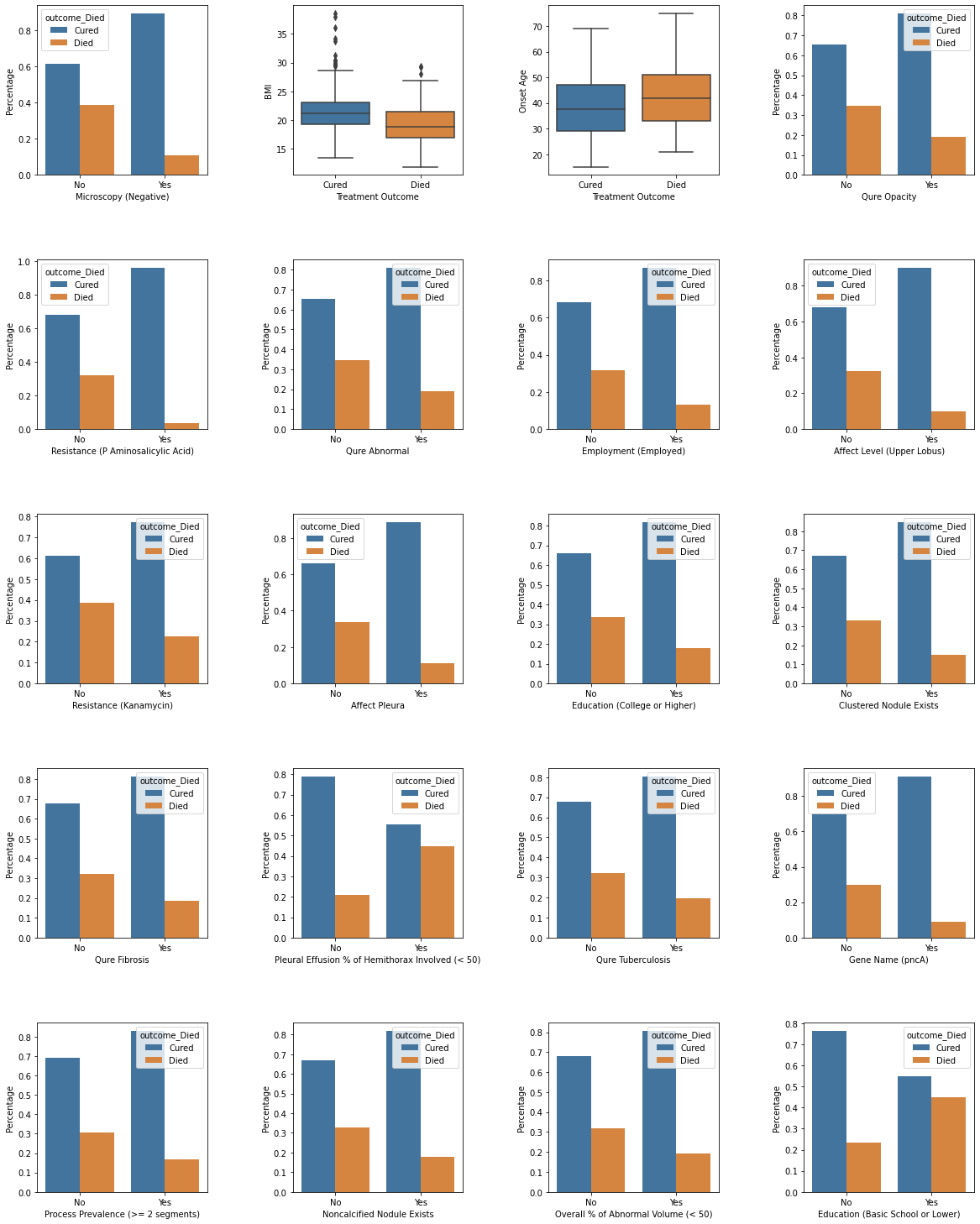

Figure S1 (c) - Top 20 Features Associated with Treatment Outcome – XDR TB

Bar charts of top 20 features from the best models stratified by drug-resistance subgroup

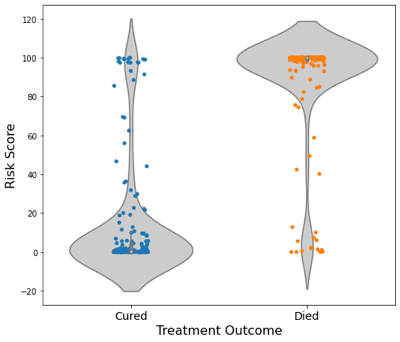

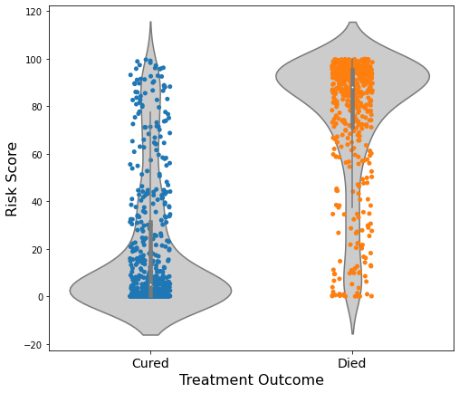

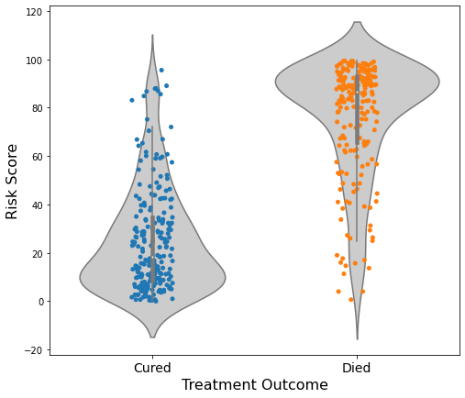

(a) DS-TB (N=299)	(b) MDR-TB (N=883)	(c) XDR-TB (N=393)

Figure S2 – Violin Plot of Risk Score

Violin plots of risk scores stratified by drug-resistance subgroup. In all three patient subgroups, the risk scores are significantly different between cured patients and dead patients (Two-sample t-test, p-value<0.0001.

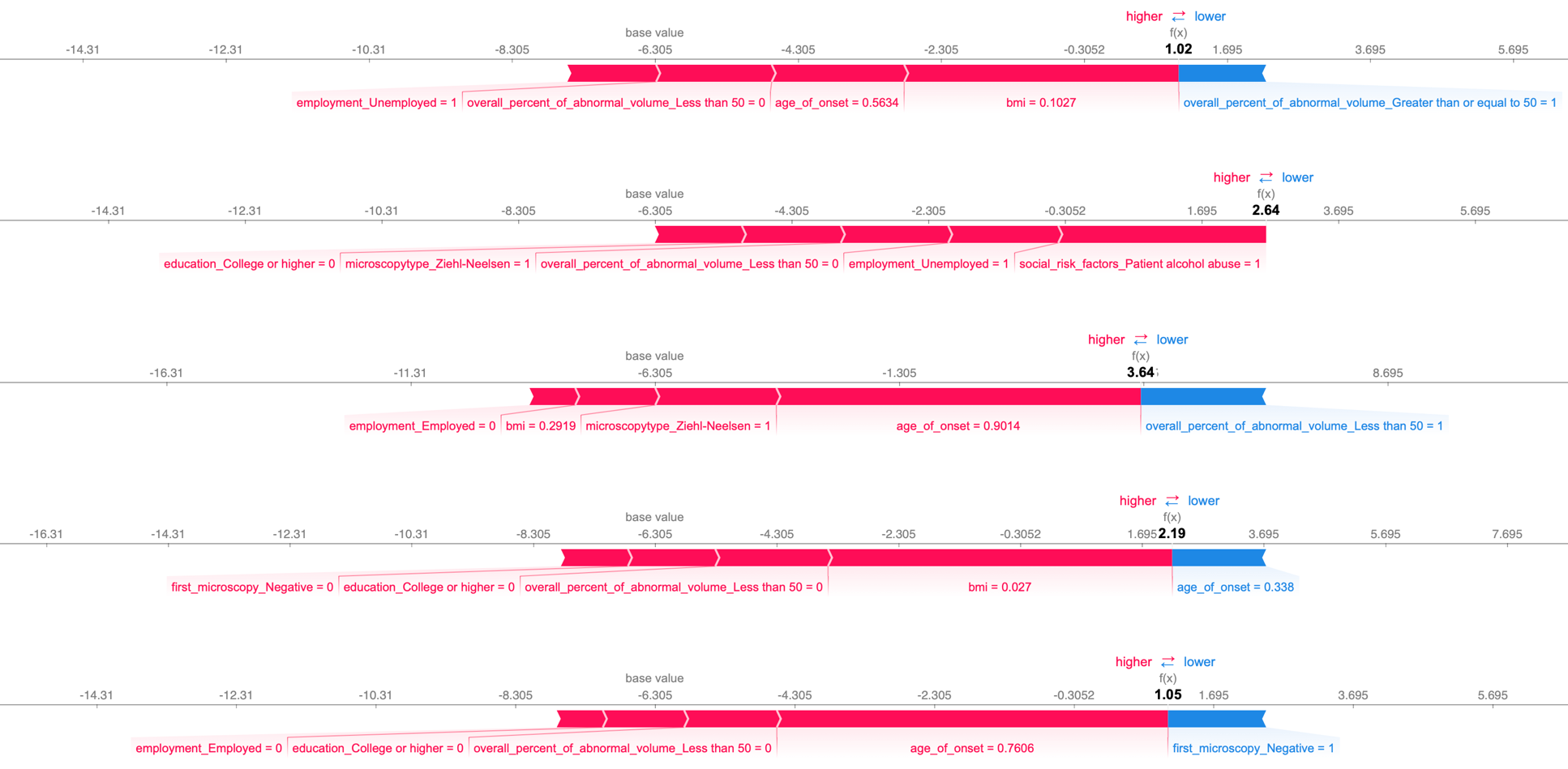

(a) Cured Patients with High Risk Scores – Sensitive TB

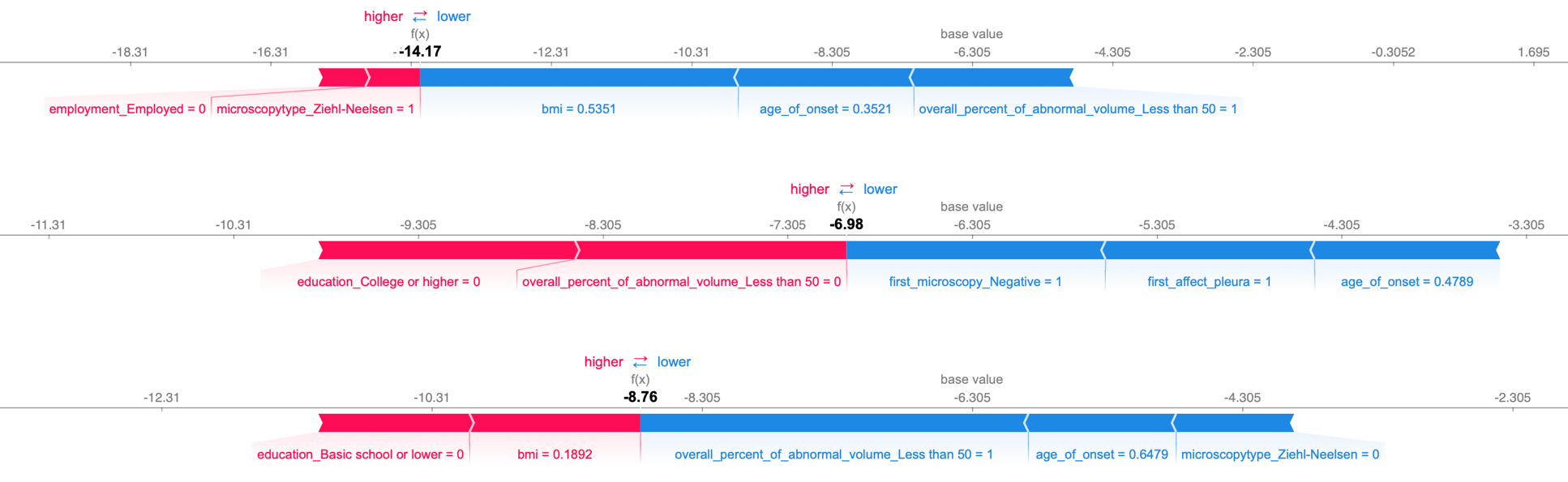

(b) Died Patients with Low Risk Scores – Sensitive TB

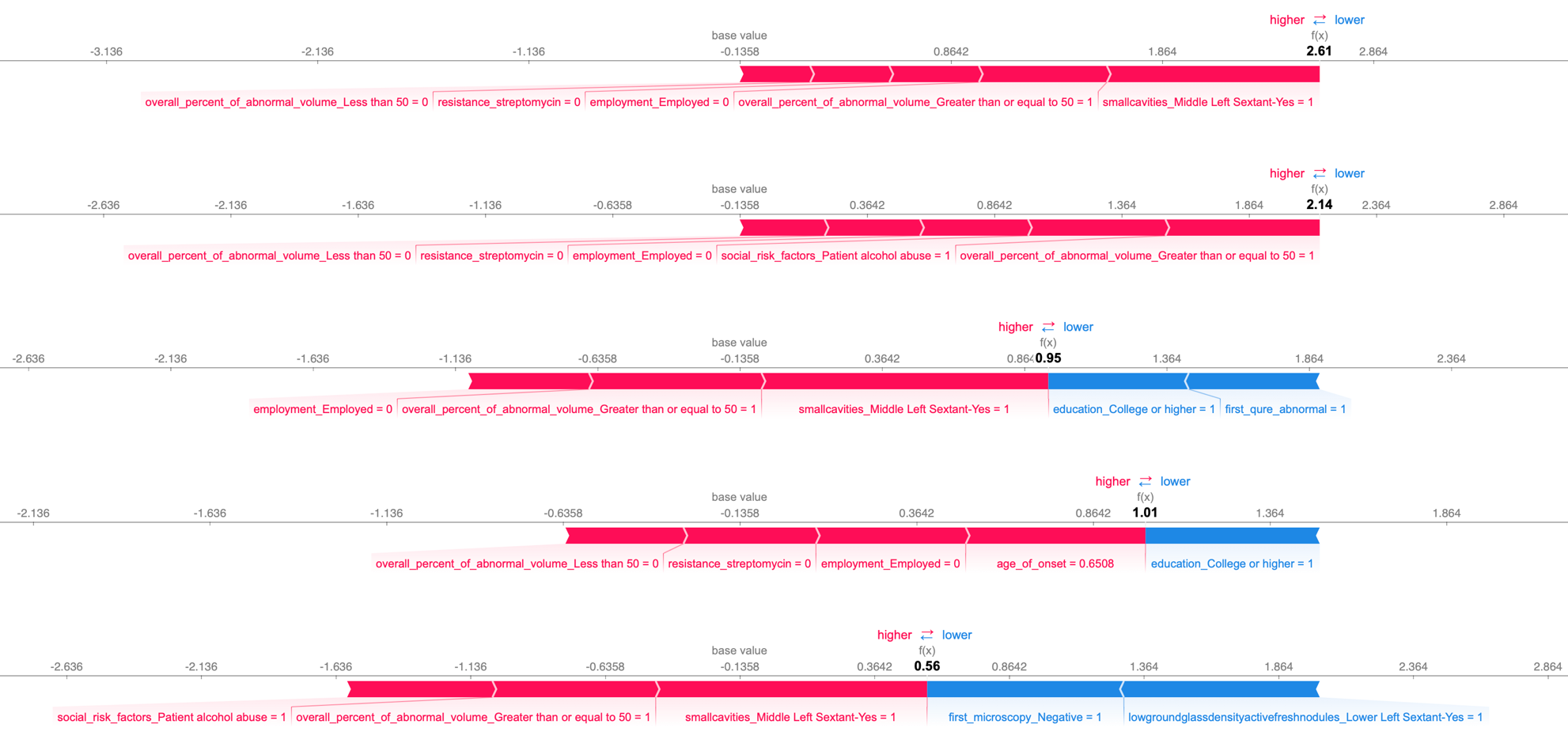

(c) Cured Patients with High Risk Scores – MDR TB

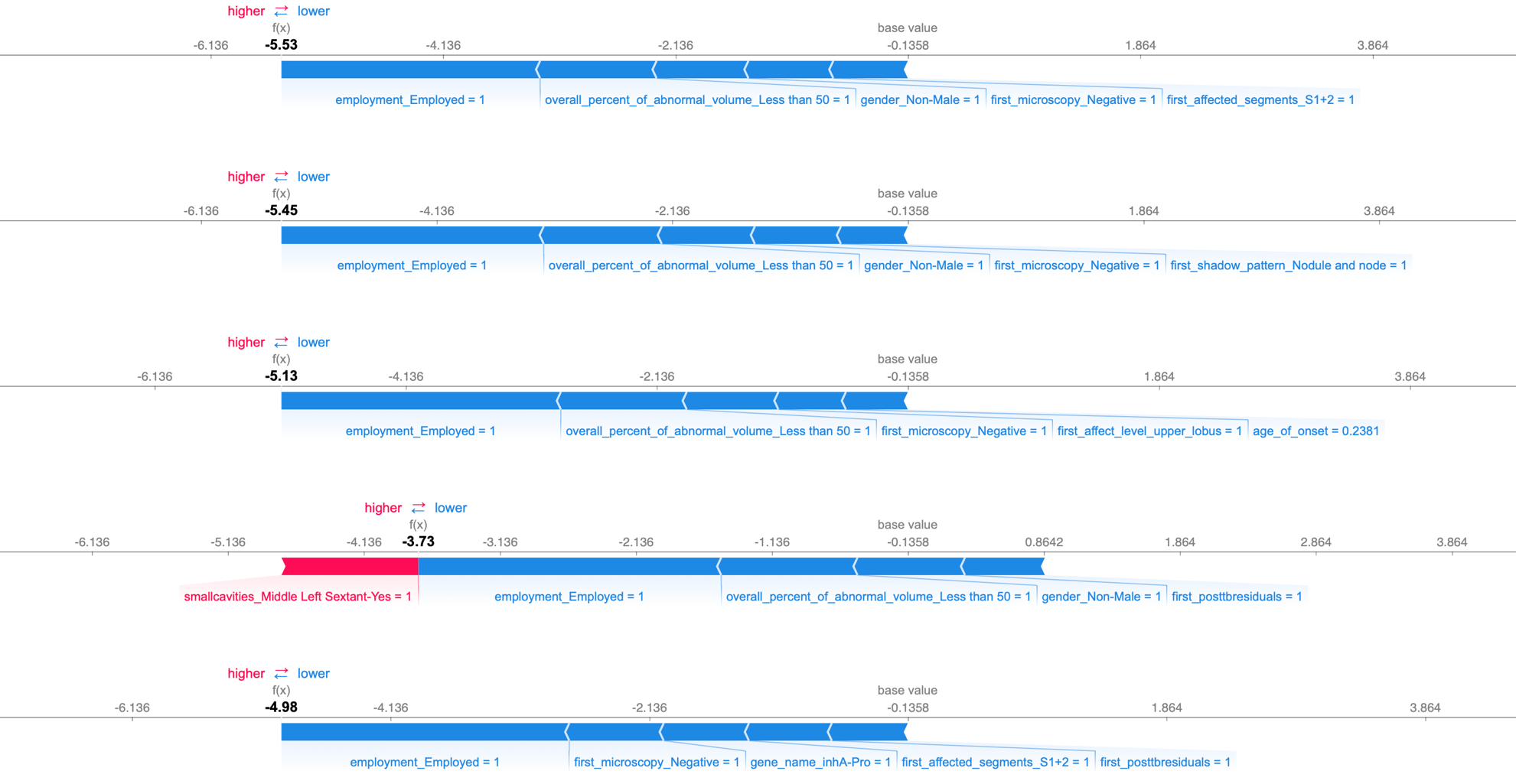

(d) Died Patients with Low Risk Scores – MDR TB

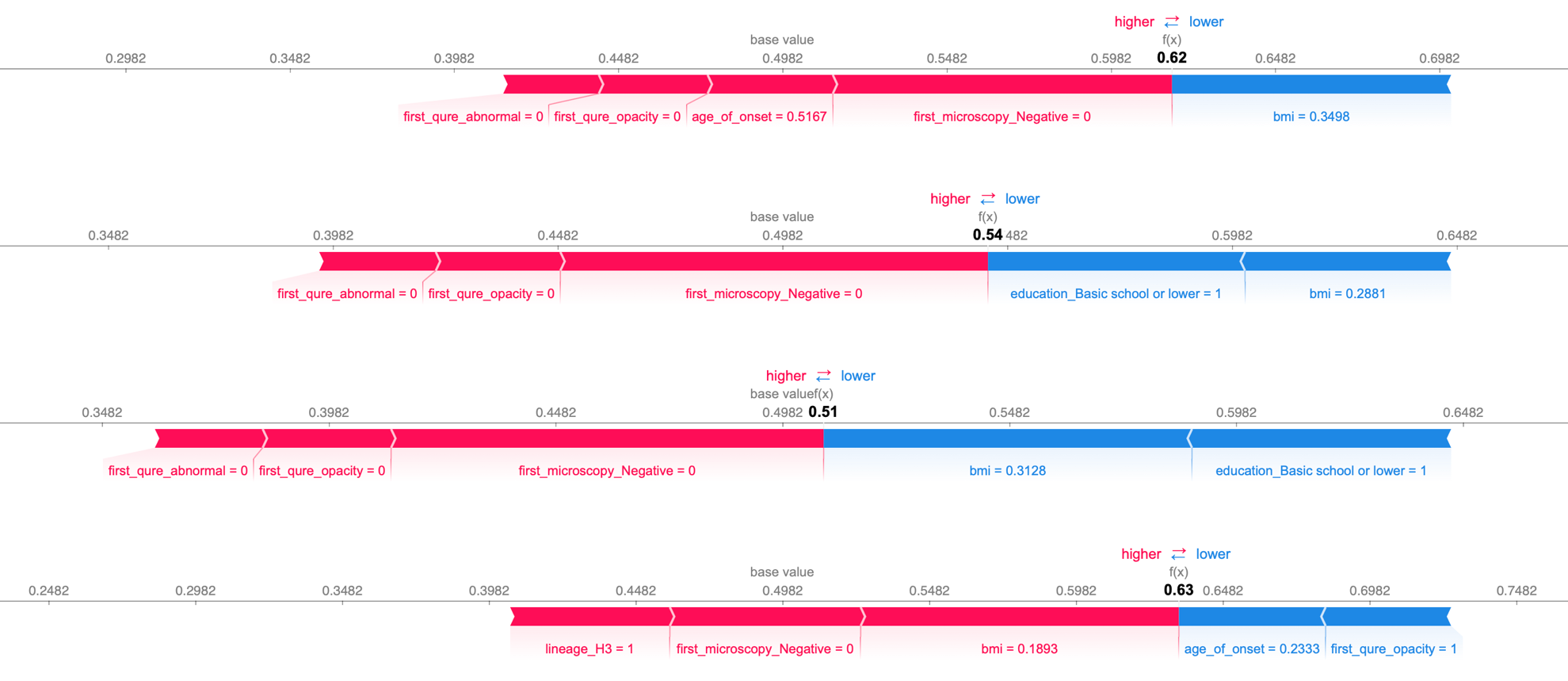

(e) Cured Patients with High Risk Scores – XDR TB

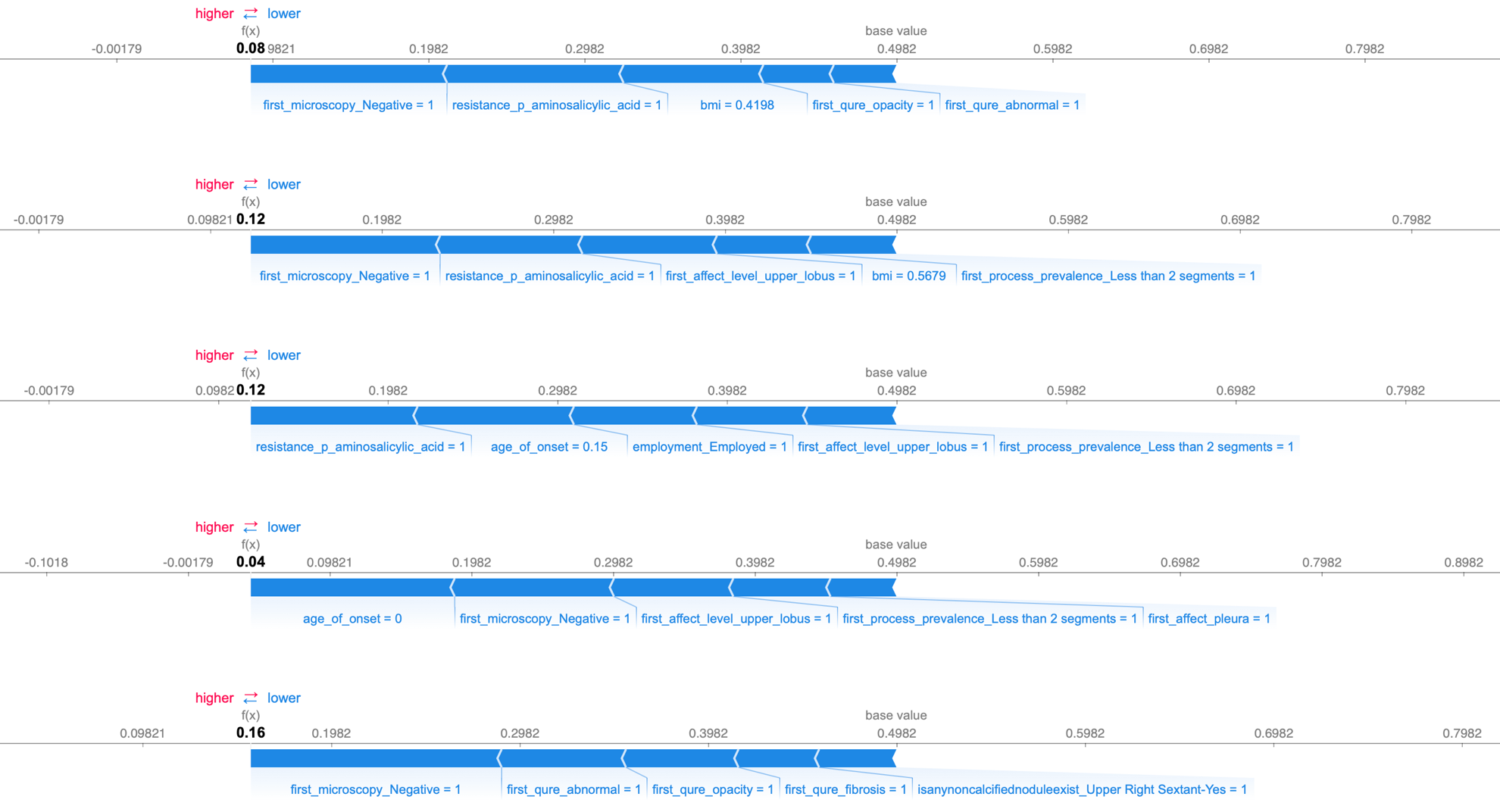

(f) Died Patients with Low Risk Scores – XDR TB

Figure S3 – SHAP Force Plots for Misclassified Patients in the Individual Testing Dataset

The SHapley Additive exPlanations (SHAP) force plots for the marginal contribution of 5 features with the highest shapley values stratified by drug-resistance subgroup. In the SHAP force plots, base value is the null model prediction (model without any features). Features in red color influence positively while features in blue color influence negatively. BMI and onset age are standardized by min-max normalization, so they range from 0 to 1. Age, BMI, negative microscopy can have a big impact as can overall abnormal on X ray but depends upon context. Sometimes the model gets it wrong if patient has weak or contrary high impact factors, but still on average the systematic modeling will provide a reasonable risk score.
